# Zero-shot extraction of seizure outcomes from clinical notes using generative pretrained transformers

**DOI:** 10.1101/2024.11.01.24316573

**Authors:** William K.S. Ojemann, Kevin Xie, Kevin Liu, Ellie Chang, Dan Roth, Brian Litt, Colin A. Ellis

## Abstract

**Purpose:** Pre-trained encoder transformer models have extracted information from unstructured clinic note text but require manual annotation for supervised fine-tuning. Large, Generative Pre-trained Transformers (GPTs) may streamline this process. In this study, we explore GPTs in zero- and few-shot learning scenarios to analyze clinical health records.

**Materials and Methods:** We prompt-engineered LLAMA2 13B to optimize performance in extracting seizure freedom from epilepsy clinic notes and compared it against zero-shot and fine-tuned Bio+ClinicalBERT models. Our evaluation encompasses different prompting paradigms, including one-word answers, elaboration-based responses, prompts with date formatting instructions, and prompts with dates in context.

**Results:** We found promising median accuracy rates in seizure freedom classification for zero-shot GPT models: one-word – 62%, elaboration – 50%, prompts with formatted dates – 62%, and prompts with dates in context – 74%. These outperform the zero-shot Clinical BERT model (25%) but fall short of the fully fine-tuned BERT model (84%). Furthermore, in sparse contexts, such as notes from general neurologists, the best performing GPT model (76%) surpasses the fine-tuned BERT model (67%) in extracting seizure freedom.

**Conclusion:** This study demonstrates the potential of GPTs in extracting clinically relevant information from unstructured EMR text, offering insights into population trends in seizure management, drug effects, risk factors, and healthcare disparities. Moreover, GPTs exhibit superiority over task-specific models in contexts with the potential to include less precise descriptions of epilepsy and seizures, highlighting their versatility. Additionally, simple prompt engineering techniques enhance model accuracy, presenting a framework for leveraging EMR data with zero clinical annotation.

**Competing Interests:** The authors declare no financial or non-financial competing interests.

## 1. Introduction

The Electronic Health Record (EHR) is a rich source of medical data, much of it in the form of unstructured text. Manual chart review is laborious and limits the scale of retrospective clinical research. Recent advances in natural language processing make automated text extraction a tractable approach to large scale medical research[1–4]. Natural language processing algorithms range in complexity from simple rules-based algorithms to cutting-edge deep learning methods. Yet even for the most advanced methods clinical text extraction can be challenging due to disease-specific terminology, variability in note style and formats, and depth of information, among other factors[5,6]. Understanding the strengths and limitations of different NLP approaches to clinical tasks is an important step towards optimizing these tools for use in clinical research.

We recently developed and validated an NLP tool for extracting clinical outcome measures in the case of people living with epilepsy. Epilepsy is one of the most common neurologic disorders, characterized by recurrent, unprovoked seizures. Among the most important outcome measures is seizure freedom. We used BERT (Bidirectional Encoder Representations from Transformers, from Google AI) language models[7], fine-tuned on manually annotated clinical notes, to extract this outcome measure with performance similar to that of human readers[4]. This approach allowed us to study the natural history and healthcare disparities in our patient population at large scale[3,8].

Yet several challenges remain to be solved. For example, this approach requires manual annotation for fine-tuning, which adds task-specific training data to a pre-trained language model. This up-front manual effort can be a barrier to rapidly generalizing the approach to new tasks or diseases and may require clinical expertise that may not always be readily available. We also have found that our algorithm performed less well on notes written by clinicians who were not specialists in epilepsy[9]. Optimizing NLP performance across a range of datasets and contexts is an important challenge for deploying these methods for large scale clinical research. Finally, our task involves temporal relations: the specific task is to determine whether the patient has had a seizure since the last office visit or within the past year, whichever is sooner. Temporal relations present a challenge for language processing algorithms and the best approach to such tasks is not yet well established[10–14].

Generative pretrained transformers (GPTs) demonstrate remarkable ability to understand text across disparate sources and domains with little or no fine-tuning on annotated datasets[15–17]. Instead of fine-tuning with manually annotated datasets, GPTs can learn through prompt engineering, providing synthetic model responses[18] and even a few examples as guidance on the desired task[17]. In a medical context, a study on extracting headache frequency from clinical notes found that a GPT2 generative model outperformed ClinicalBERT[19]. Still, fully fine-tuned encoder models can outperform un-finetuned GPTs in specific tasks; for example, Guo et al. found that fully-finetuned BERT models outperformed various GPTs in most tests within a finance domain[20].

In this study, we aimed to test the capabilities of publicly available GPTs for extracting clinical outcomes from free text notes without manual annotation. We compared the performance of Llama2 GPT models with prompt engineering against the benchmark of our fine-tuned BERT model. We compared several prompt engineering strategies, focused on addressing the temporal reasoning aspect of our task. We also tested how well this approach generalized to notes written by nonspecialists, a particular challenge for the BERT method. Finally, we sought to replicate a previous clinical application of our model, to determine whether we could have achieved the same results without manual annotation effort if we had used Llama2 instead of BERT in that study.

## 2. Methods

This research was approved by the Institutional Review Board of the University of Pennsylvania.

### 2.1 Data collection

#### 2.1.1 Annotated datasets for performance evaluation

Our source dataset for fine-tuning and validation consisted of 78,844 progress notes for patients with epilepsy who visited the University of Pennsylvania Health System between 2015 – 2018[4]. We have previously annotated, in triplicate, 1000 notes written by epileptologists. Annotators were asked to determine if the note suggested a patient was seizure free or having recent seizures, the patient’s seizure frequency, and/or the date of their last seizure. We defined a “seizure free” visit as one where the patient did not have seizures since their last visit, or within the past year, whichever was more recent. We then merged the triple-annotated documents through majority voting, with manual adjudication of disagreements. 700 of these annotated ground-truth documents became the training set, and 300 became the validation set.

We repeated this annotation process for 100 notes written by neurologists who were not specialized in the management of epilepsy, and 100 notes from all other clinical providers – generalists. These 200 notes were annotated by two authors (K.X. and C.A.E) under the same annotation protocol; annotations were merged with manual adjudication in the event of disagreement[9].

#### 2.1.2 Dataset for demographics reproduction

Our source dataset for replicating our findings on demographic disparities in outcome measures consisted of electronic health records from 25,612 unique patients who had seen an epileptologist at the University of Pennsylvania Health System between 2005 and 2022. These records included the full text of their progress and discharge notes, prescription records, and patient metadata.

### 2.2 Bio+ClinicalBERT model development

We recently developed and validated an NLP algorithm that classifies clinic notes as seizure-free or having recent seizures[4]. Briefly, we used Bio+Clinical_BERT a publicly available transformer language model from Google AI[21], on 700 manually annotated epileptologist notes. Model predictions were repeated five times using different seeds, and final classification of each note was determined by plurality voting of the five outputs. Our goal in this study was to test several zero- and few-shot prompt engineering strategies and compare their performance to this benchmark model.

### 2.3 GPT architecture and system prompt

We performed our experiments on the EHR using Llama2 13B, an open-source GPT model from META Gen AI[22]. Llama2 13B can accept a context window of 4096 tokens. Parameters for Llama2 13B were gathered from chat fine-tuned versions stored on huggingface.com (https://huggingface.co/meta-llama/Llama-2-13b-chat-hf).

We asked Llama2 to answer the following question: “given a note, has the patient been seizure free in the past 12 months,” with three possible outputs: yes, no, or unknown. We modified the system prompt to say that the model should respond as a board-certified neurologist, described an epileptic seizure, and asked the model to draw on information from the note to answer the question of seizure freedom (see **Supplementary Material** for the system prompt text).

### 2.4 Prompt engineering experiments

We conducted five prompt engineering experiments (**Table 1**): (1) One-word, (2) Elaboration, (3) Date formatting, (4) Date-in-context, (5) Instruction fine-tuning. For each experiment we used the python *time* package to calculate the elapsed time for prediction generation. Full prompt text for each experiment is available in the **Supplementary Material**.

**Table 1.**
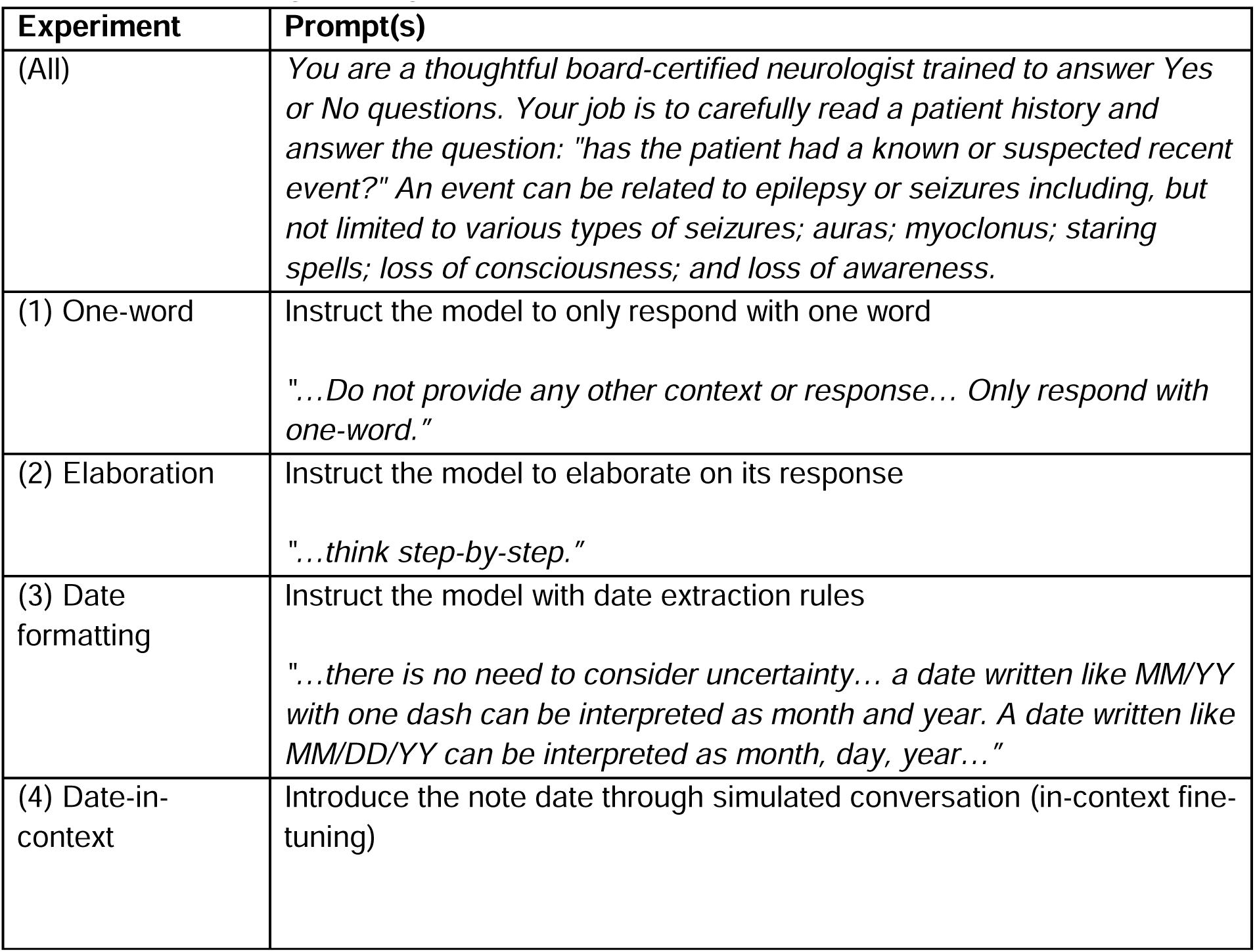
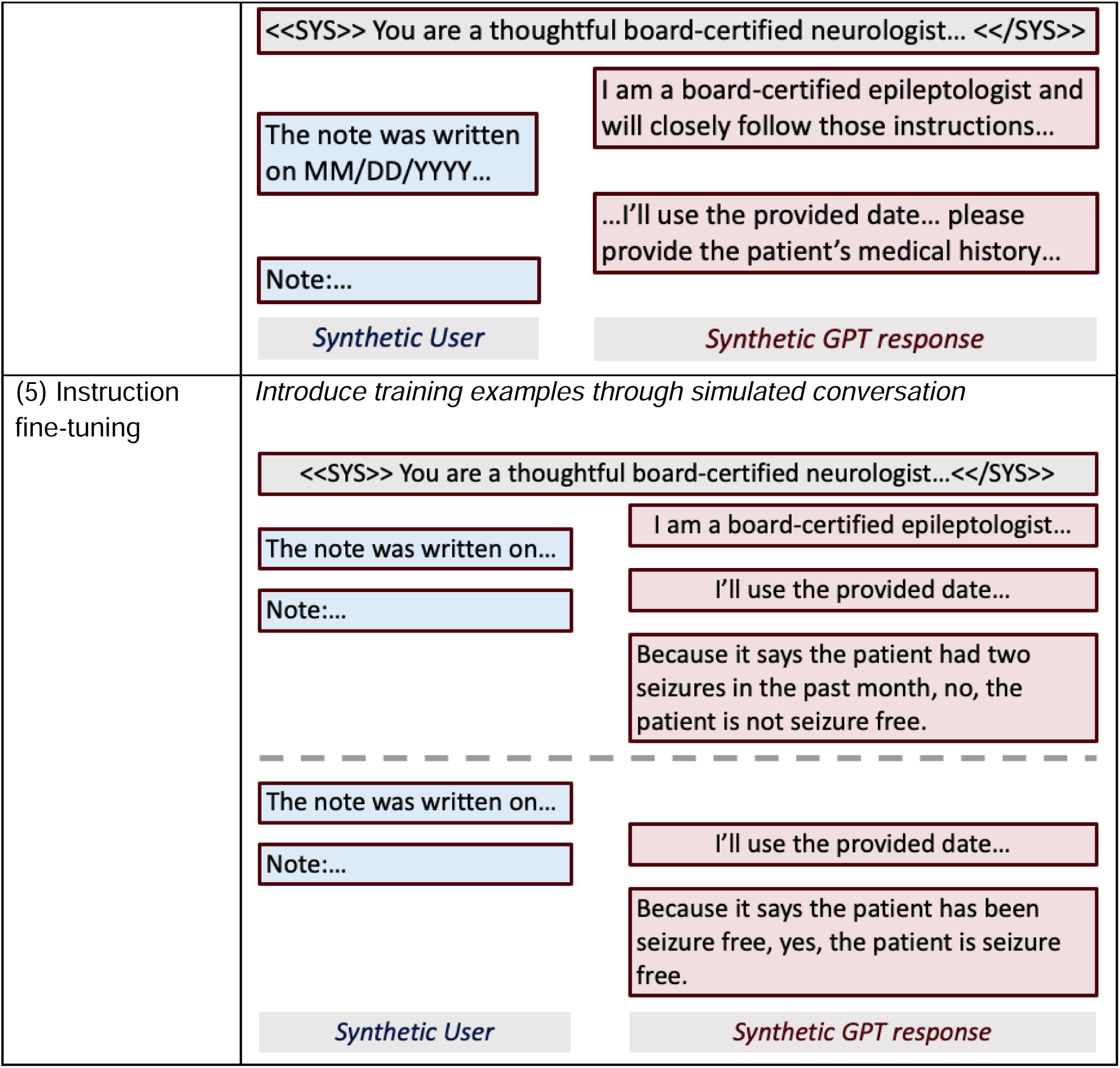
Prompt engineering experiments.

#### 2.4.1 Experiment 1: One-word seizure freedom extraction

The prompt instructed the model to respond with a single word: yes, no, or unclear. The prompt instructed the model not to provide any other context or response in addition to your one-word response.

#### 2.4.2 Experiment 2: Elaboration

Because recent work has demonstrated that LLMs perform better in classification tasks when given the chance to reason[23], we next constructed a new system prompt that guided the LLM to provide rationale in addition to its one-word response. To facilitate reasoning in the response, we appended the instruction to “think step-by-step” at the end of the prompt so the model would explain the rationale behind its answers.

#### 2.4.3 Experiment 3: Date formatting

This prompt included guidelines for parsing the MM/DD/YYYY date format. It also included a clearer definition of epileptic seizures along with specific, common boundary cases in our training dataset to specify that surgeries and taking medicines are not epileptic events, and logical rules for response generation to instruct the model to output “yes” in cases of seizures.

#### 2.4.4 Experiment 4: Date-in-context

In this experiment, after the system prompt we synthesized a model response acknowledging the instructions. Then we provided a second prompt containing the date the note was written, followed by another synthesized model response acknowledging the date. Finally, we provided the medical note for classification.

#### 2.4.5 Experiment 5: Instruction Fine-tuning

Instruction fine-tuning provides training data to GPTs in the form of a conversation history. We applied the date-in-context method with instruction fine-tuning, using 3, 5, and 8 training samples. For each sample size, we conducted 5 independent iterations of random sampling without replacement from the training corpus, permitting potential overlap of notes between iterations. The expected model response to each training sample was inserted into the chat history as part of the synthetic conversation history.

#### 2.4.6 Extracting output classifications

When the Llama2 model outputs were longer than one-word (experiments 2-5), we used a fine-tuned Bio+ClinicalBERT model to classify the outputs into the desired output categories (Yes, No, Unclear). First, in each experiment we manually classified 600 of the Llama2 outputs into these trinary categories. We then used 70% of the 600 classified outputs for training the Bio+ClinicalBERT classification model, and 30% for testing. This method achieved 99% accuracy and the results are not presented separately below.

### 2.5 Performance evaluation and benchmarks

For each experiment, we compared accuracy in classifying seizure freedom on our test set of 300 annotated epileptologist notes against three benchmarks: (1) Bio+ClinicalBERT without fine-tuning (zero-shot); (2) Fine-tuned Bio+ClincialBERT; (3) a majority class classifier, i.e., the accuracy a model would achieve if it predicted the most common classification for all samples. We also compared the LLM’s performance with 0, 3, 5, and 8 instruction fine-tuning samples to the fully fine-tuned Bio+ClinicalBERT model. To compare Llama2 against Bio+ClinicalBERT fine-tuned with different amounts of training data, we fine-tuned with varying amounts of training data from 0 to 700 notes in 10% (70 note) increments.

### 2.6 Generalization to novel clinical contexts

To evaluate the generalizability of our LLM to other note types, we compared the classification accuracy for seizure freedom of our prompt-engineered LLM in a zero-shot setting to the previously validated Bio+ClinicalBERT models. We also compared the accuracy of all models against an oracle majority class classifier for each testing dataset – epileptologist, neurologist, and generalist notes.

### 2.7 Analysis of disparities in seizure outcomes

In a recent study of the same dataset used here, we investigated health disparities in seizure outcomes between demographic groups[3]. Here we tested the hypothesis that a zero-shot approach using Llama2 and prompt engineering would have been able to detect those same disparities and reach the same conclusions without a fine-tuned model. We followed the same analysis methods as the previous study[3]. Summarizing the analysis, the demographic variables were race, sex, ethnicity, age, median zip code income, and insurance type. We tested for differences between demographic groups in terms of their likelihood of seizure-free office visits, using univariable and multivariable linear mixed-effects models to account for repeated measures and variations in time between visits. Here we used the output of the Llama2 date-in-context prompt (experiment 4) as the source of seizure outcome data, in place of the Bio+ClinicalBERT generated data in our previous study and compared the two sets of results.

### 2.8 Statistical analysis

For each Bio+ClinicalBERT model, we repeated the fine-tuning step five times using different seeds to generate estimate the variance and confidence intervals for performance and inference time. For the experiments using Llama2, we found that the greedy token selection algorithm performed better than stochastic algorithms that allow for some randomness in token selection (see **Supplementary Material**). Therefore, we could not introduce variability in outputs using different seeds. Instead, we calculated bootstrap confidence intervals, using 15 bootstraps of 100 samples of the testing dataset in each analysis. We tested for differences in performance and inference time between models and prompts using non-parametric, two-tailed Mann-Whitney U tests. We tested for differences in model performance against majority-class classifiers using non-parametric, one-tailed Wilcoxon signed-rank tests. P-values were corrected for multiple comparisons using a Benjamini-Hochberg false discovery rate of 0.05[24]. Distributions of values are reported as 50^th^ [25^th^, 75^th^] percentiles.

## 3. Results

### 3.1 Experiments 1-4: Zero-shot prompt engineering

Among the first four prompt engineering experiments (**Figure 1**), the date-in-context prompt (experiment 4) performed the best (median accuracy 74%, IQR [73%, 80%]) compared to one-word (62% [58%, 65%]), elaboration (50% [48%, 54%]), and date formatting (62% [62%, 66%]). All Llama2 prompts significantly outperformed the Bio+ClinicalBERT zero-shot model (25% [23%, 29%]), and performed significantly worse than the fully fine-tuned Bio+ClinicalBERT model (84% [83%, 84%]) (Mann-Whitney U: p < 0.01). All differences between prompt engineering methods were significantly different (Mann-Whitney U: p < 0.001) except for date formatting vs. one-word (Mann-Whitney U: p = 0.23). Only the date-in-context model outperformed the majority class benchmark (Wilcoxon: p < 0.001) while the one-word (Wilcoxon: p = 0.60) and date formatting (Wilcoxon: p = 0.22) models did not outperform the majority class benchmark, and the elaboration model underperformed the majority class benchmark (Wilcoxon: p < 0.001).

**Figure 1.**
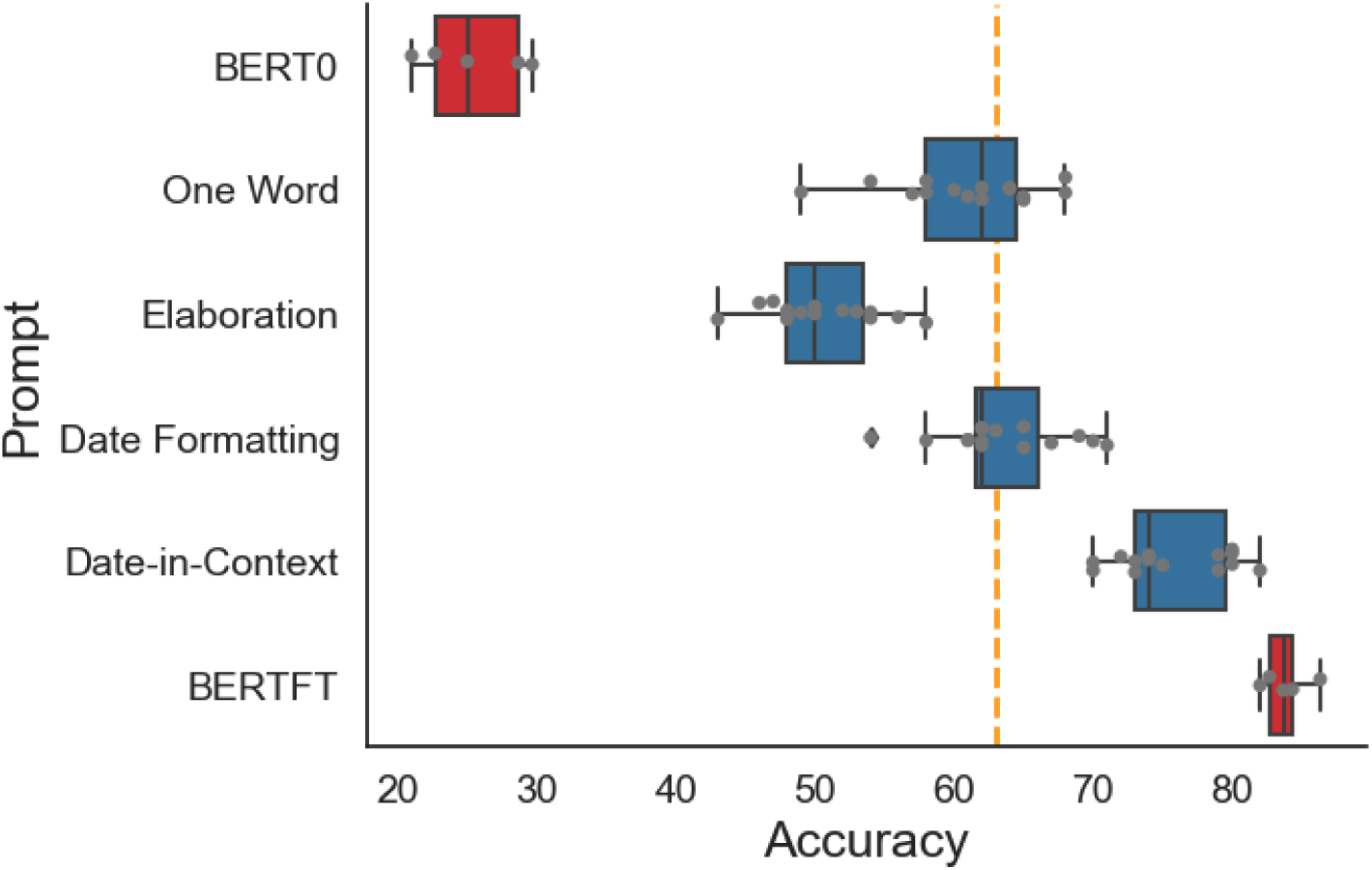
Zero-shot prompt engineering. Results of four different prompt engineering strategies using Llama2 (blue), compared to benchmarks of Bio+ClinicalBERT without fine-tuning (BERT0), Bio+ClinicalBERT with fine-tuning (BERTFT), and a majority class classification (yellow dashed line).

### 3.2 Experiment 5: Instruction fine-tuning

Introducing training examples to the date-in-context prompt reduced performance on the classification task (**Figure 2**). The zero-shot model (median accuracy 76%, 95% CI [74%, 81%]) significantly outperformed the 3-sample (68% [65%, 70%], Mann-Whitney U: p = 0.0025) and 8-sample instruction fine-tuning models (61% [58%, 66%], Mann-Whitney U: p = 0.0012), while there was no detected difference between the zero-shot and 5-sample instruction fine-tuning models (76% [72%, 76%]) (Mann-Whitney U: p = 0.54).

**Figure 2.**
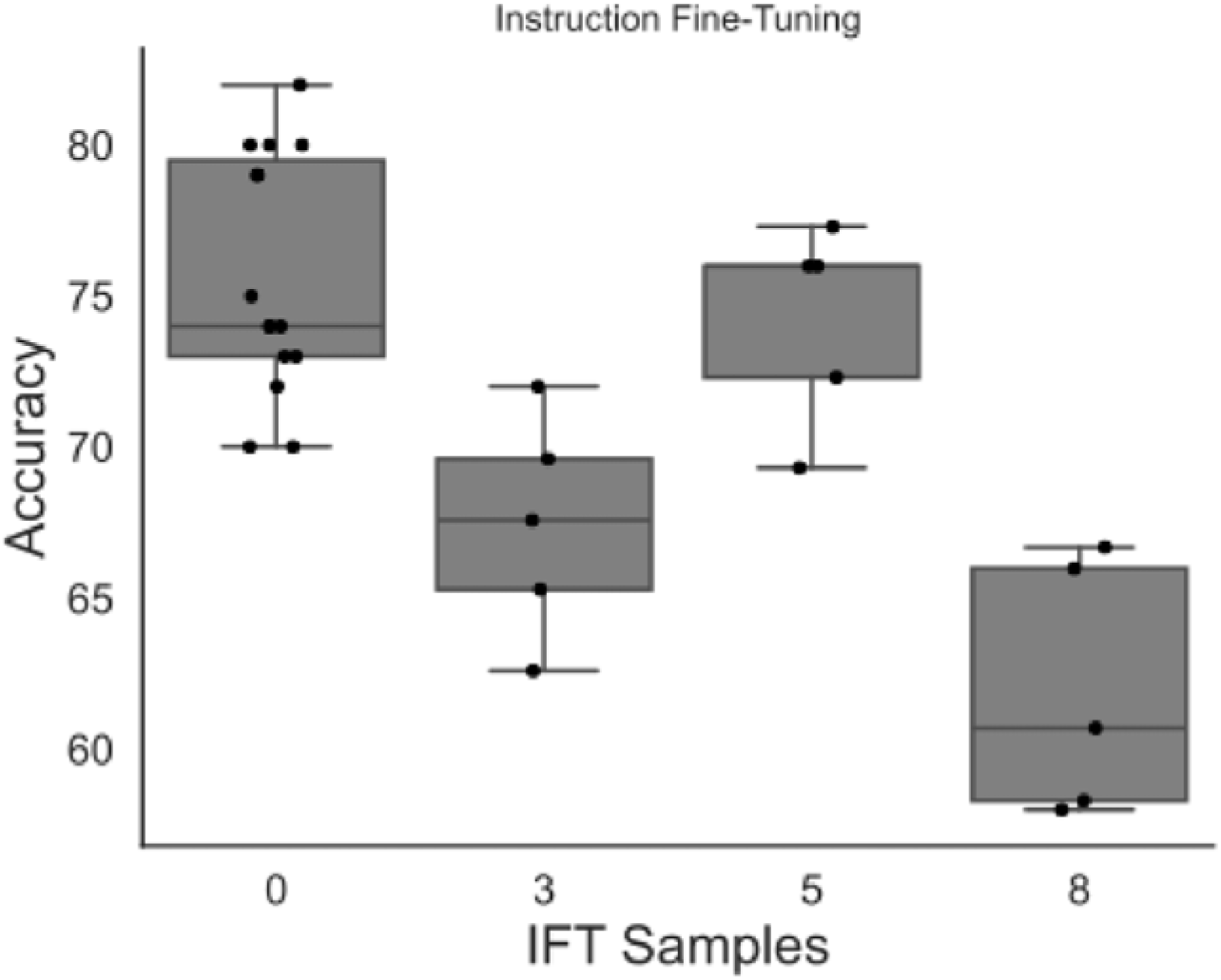
Results of instruction fine-tuning. Accuracy of the Llama2 model with date-in-context prompt, adding different numbers of training examples to the prompt through instruction fine-tuning. Abbreviation: IFT = Instruction fine-tuning

### 3.3 Comparative evaluation of fine-tuning sample size

Having found that the best-performing zero-shot Llama2 model (date-in-context prompt, experiment 4) was less accurate than the benchmark Bio+ClinicalBERT model fine-tuned on 700 training samples, but more accurate than zero-shot Bio+ClinicalBERT, we next asked how many training samples were needed for Bio+ClinicalBERT to match and exceed zero-shot Llama2. We found that Bio+ClinicalBERT was inferior to zero-shot Llama2 with 140 or fewer training samples and did not achieve significantly better results than Llama2 until 420 training samples (**Figure 3**).

**Figure 3.**
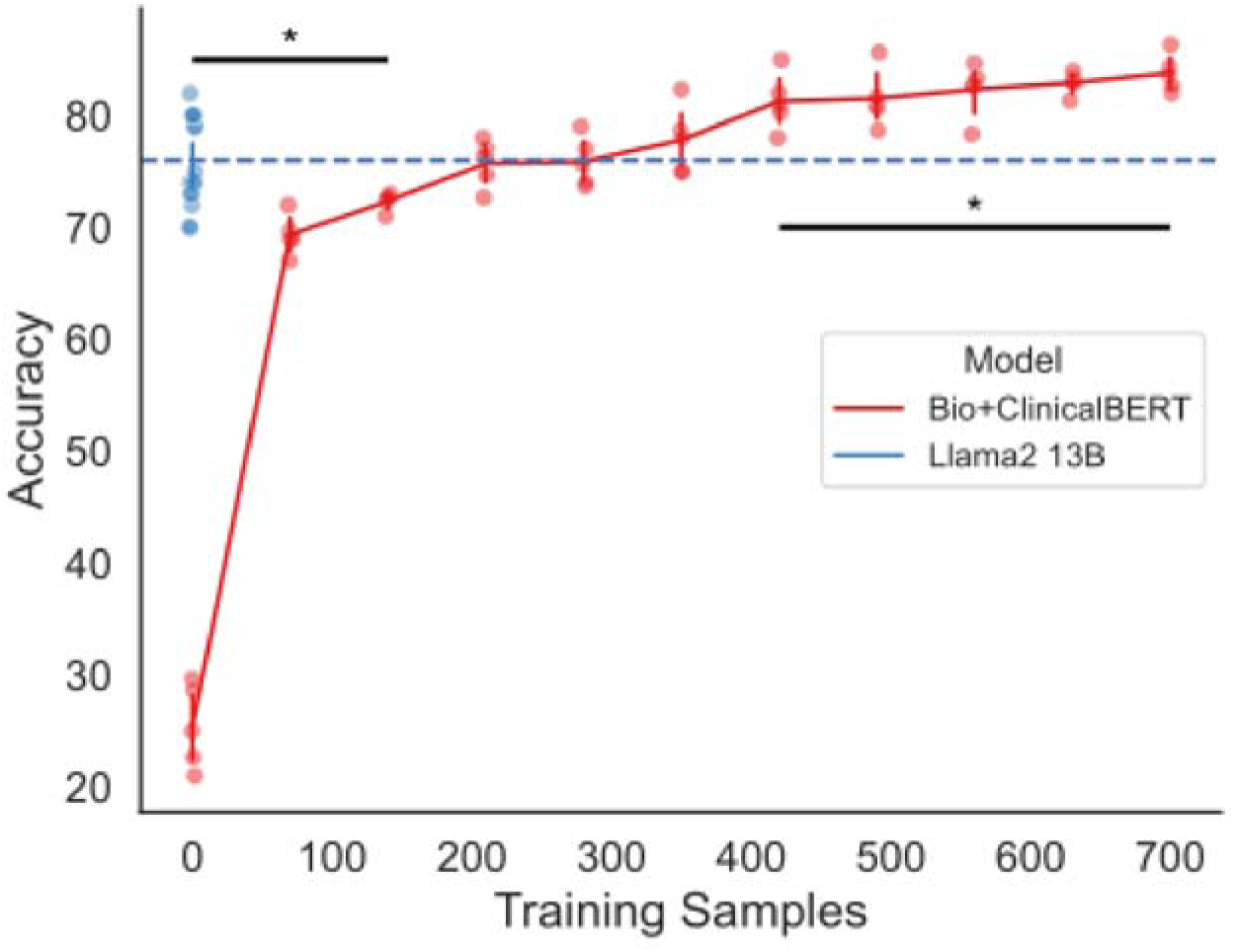
Training data required for fine-tuning Bio+ClinicalBERT. Results of increasing training data used to fine-tune Bio+ClinicalBERT (red), in increments of 10% (70 notes), compared to zero-shot Llama2 using date-in-context prompt engineering. Blue dashed line shows the median Llama2 performance for reference. Bars above (below) accuracy distributions indicate significantly higher (lower) performance when comparing Llama2 to Bio+ClinicalBERT.

### 3.4 Model generalizability to non-specialist notes

Llama2 date-in-context significantly outperformed the generalist (35%) and neurologist (47%) majority class classifiers (Wilcoxon: p < 0.001). For notes written by non-neurologists, there was no significant difference in accuracy between Llama2 (49%, [47%, 52%]) and the fine-tuned Bio+ClinicalBERT (52%, [51%, 57%], Mann-Whitney U: p = 0.052). For notes written by neurologists who were not specialists in epilepsy, Llama2 (76%, [74%, 81%]) significantly outperformed Bio+ClinicalBERT (67%, [65%, 70%], Mann-Whitney U: p = 0.0051). There was no significant difference between the performance of Llama2 on neurologist notes and epileptologist notes (Mann-Whitney U: p = 0.62) (**Figure 4**).

**Figure 4.**
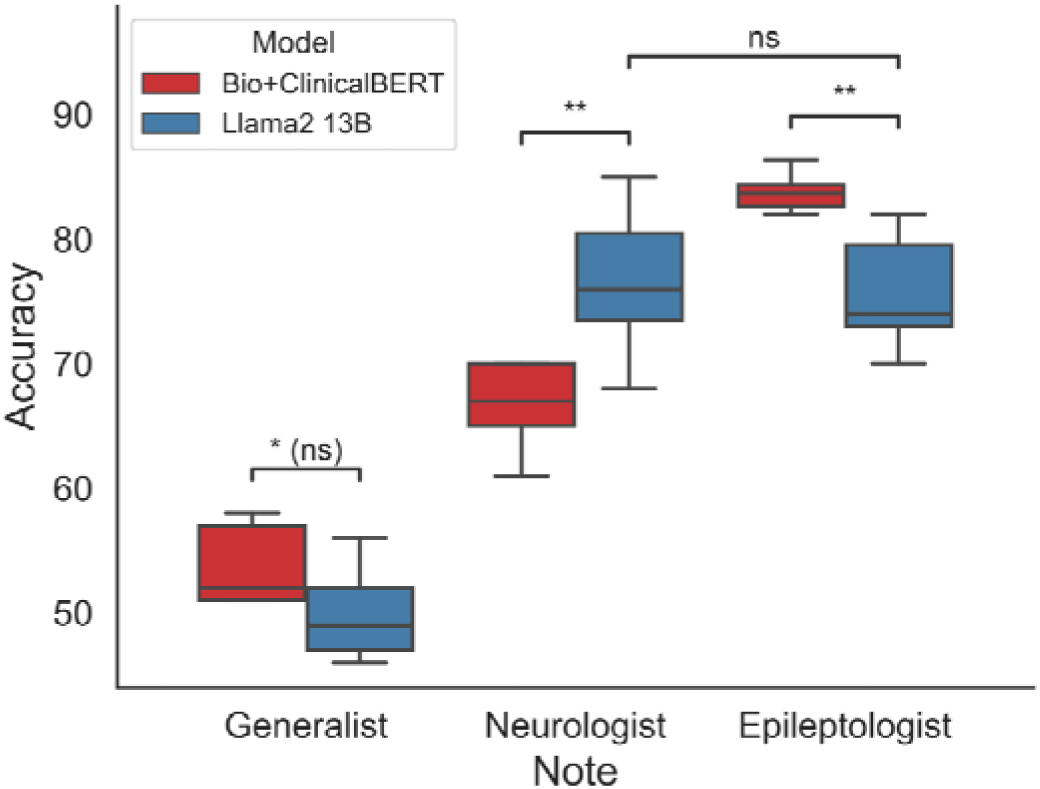
Model generalizability to nonspecialist notes. Performance of Bio+ClinicalBERT (red) and zero-shot Llama2 with date-in-context prompt engineering (red) on notes written by three different groups of authors: epilepsy specialists (epileptologists), neurologists who were not specialists in epilepsy, and non-neurologists. The Bio+ClinicalBERT model was fine-tuned on notes written by epileptologists.

### 3.5 Clinical application of different NLP methods

We reproduced a previously published analysis of health disparities in seizure outcomes of 84,675 notes from 25,612 patients using two different NLP methods[3]. Llama2 with date-in-context prompts preserved 91% (10/11) of the significant findings previously detected by the Bio+ClinicalBERT model in both univariate and multivariate analyses (**Figure 5**). The only discrepancies (i.e., instances where one model would have indicated a significant effect while the other model did not) were for female sex.

**Figure 5:**
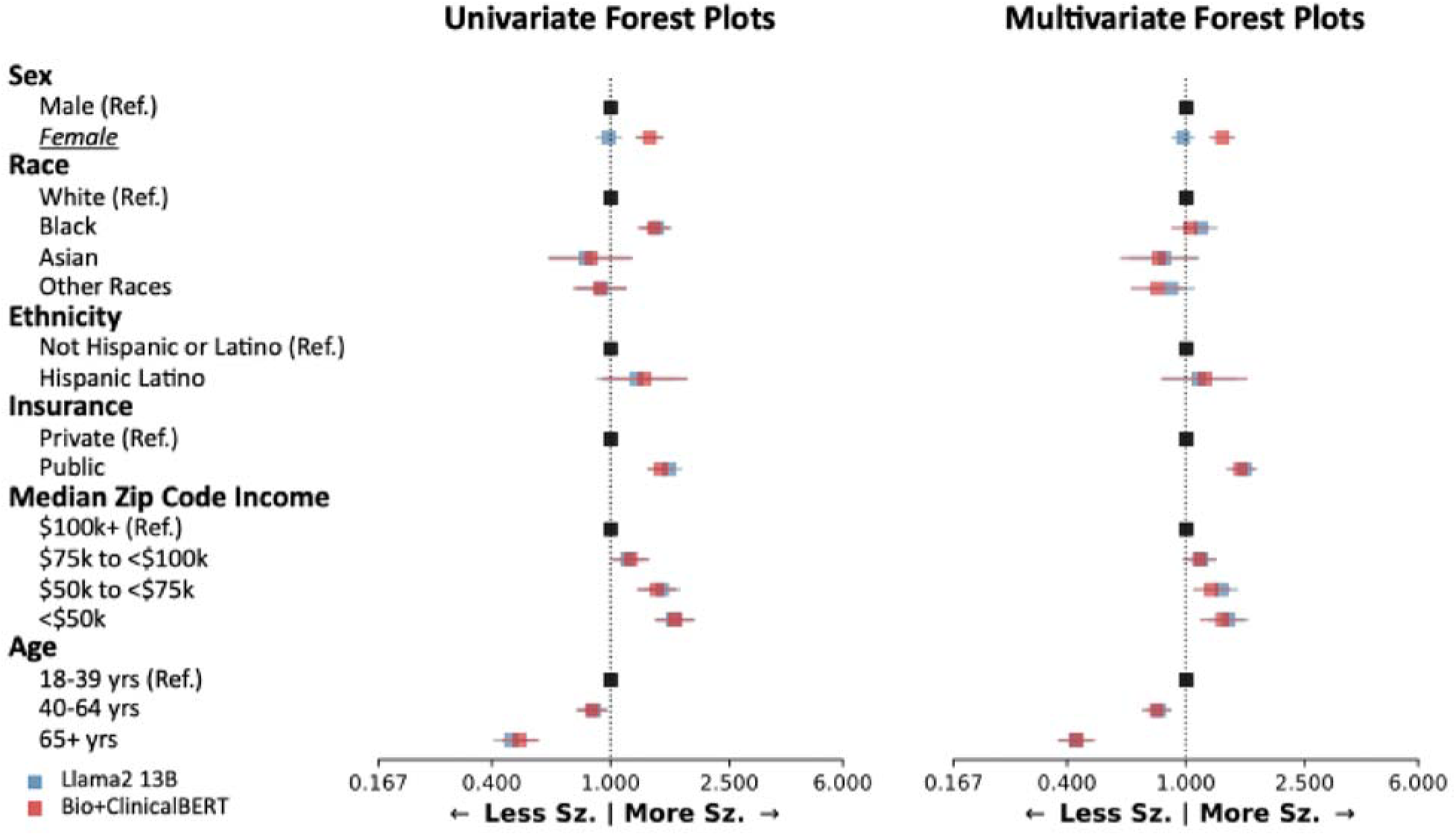
Clinical application of different NLP methods. Analysis of demographic disparities in seizure outcomes, as generated by Bio+ClinicalBERT (red) and Llama2 (blue). The left forest plot shows univariable analysis and the right forest plot shows multivariable analysis. Forest plots show odds ratios relative to reference groups. Groups with discrepant findings between Llama2 and Bio+ClinicalBERT are underlined and italicized. Abbreviations: Sz = seizure.

### 3.6 Inference times

The time to interpret 300 clinical notes was longer for all Llama2 models than the Bio+ClinicalBERT model (**Table 2**). Between the different Llama2 prompt experiments, differences in inference time were not fully attributable to prompt length: for example, the date-in-context prompt (experiment 4) had the longest prompt length, but two other models had longer inference times. When examining the relationship between inference time and instruction fine-tuning samples, inference times increased with more training examples in the prompt.

**Table 2.** Inference times.

| Experiment | Model/Prompt | Prompt length (characters) | Inference time in seconds, median [25%, 75%] |
| --- | --- | --- | --- |
| Benchmark | Bio+ClinicalBERT | n/a | 23 [22, 29] |
| Exp. 1 | One-word | 804 | 866 [865, 868] |
| Exp. 2 | Reasoning | 867 | 3574 [3571, 3575] |
| Exp. 3 | Temporal reasoning | 1573 | 5225 [5224, 5226] |
| Exp. 4 | Date-in-context | 1650 | 3445 [3442, 3445] |
| Exp. 5 | Instruction fine-tuning |  |  |
|  |  | 3 samples | 3442 [3352, 3793] |
|  |  | 5 samples | 4356 [4254, 5097] |
|  |  | 8 samples | 6924 [6672, 7186] |

## 4. Discussion

This study extracting seizure outcomes from clinical notes using NLP had several important findings. First, different prompt engineering strategies of a zero-shot Llama2 language model led to substantial changes in performance, though none achieved the benchmark performance of a fine-tuned Bio+ClinicalBERT model. Second, adding training data to the Llama2 model through instruction fine-tuning did not improve its performance. Third, the fine-tuned Bio+ClinicalBERT model required substantial training data—420 training samples—to exceed the performance of the best-performing zero-shot Llama2 model. Fourth, the best Llama2 model also performed better on notes written by nonexperts, suggesting better potential generalizability than a Bio+ClinicalBERT model fine-tuned on notes written by experts. Finally, we reproduced a clinical analysis of healthcare disparities in 84,675 notes and found similar results using both NLP methods.

Extracting data from clinical text is an important step forward for medical research. Unresolved questions include which methods and models work best for which kinds of tasks, and how much manual annotation is required for model fine-tuning. Generally, GPT models have the ability to perform well on a variety of tasks without domain-specific fine-tuning[15–17]. Our findings demonstrate that GPTs can perform the complex text extraction required to label notes for seizure freedom without any parameter fine-tuning or manually labeled data. However, consistent with literature in other domains[20], our work suggests that fine-tuning a smaller, encoder-only model with training data can eventually exceed the performance of a zero-shot model with prompt engineering. While GPT models can also be fine-tuned (i.e., parameter weights can be updated), but this requires a significant investment in computational and annotation resources.

Previous studies have compared versions of BERT and GPT language models for clinical text mining and have found similar patterns in performance. For example, Gema et al. compared fully-finetuned encoder models against Llama variants, and found that, on average, the finetuned encoder models typically outperformed the baseline Llama and clinically pretrained Llama models on outcome prediction datasets; the decoder models were able to match or outperform the encoder models only after the addition of specialized finetuning techniques[25]. Likewise, Lilli et al. found that finetuned BERT variants had largely comparable or better precision, accuracy, and F-scores to zero-shot Llama2 7B and Mixtral 8×7b when identifying the presence of metastatic tumors in Italian EHRs[26]. Wang et al. 2024 compared a finetuned ClinicalBERT to finetuned Llama models and found the latter to be better at classifying diagnosis-related groups[27]. In our study, we posit that our fine-tuned Bio+ClinicalBERT model outperformed the best zero-shot Llama2 model likely due to the nature of the temporal reasoning task, which we discuss further below, and due to the broad domain knowledge used during training[20].

The prompt engineering strategies that we tested here have been studied before in other contexts, and we evaluated combinations of prompting strategies to maximize performance of Llama2 on our seizure freedom extraction task. The prompt we test to facilitate elaboration in experiment 2 stems from Kojima et al 2022, which suggests simply instructing an LLM to "think step by step" can unlock zero-shot reasoning abilities in logical problem-solving[23]. This method also generates more nuanced responses, which we found to qualitatively allow for more interpretability. The “date-in-context” strategy in experiment 4 was adapted from Wei et al., which demonstrated significant improvements in model reasoning abilities with intermediate reasoning steps[18]. In our prompt, we introduced the note date – a key piece of information for our seizure freedom task – through this finetuning method and similarly saw a jump in performance. Instruction fine-tuning, as popularized by Brown et al., allows a model to learn tasks by observing examples provided as chat history[17]. There are multiple potential explanations for the poor performance of our instruction fine-tuning experiments. It has been shown that instruction fine-tuning teaches the semantic format of the tasks rather than introducing new reasoning or domain knowledge, which we see through decreased inference times as we provide synthetic GPT responses[28]. Further, the order of instruction fine-tuning samples has been shown to be critical for increasing performance, and, since we did not have the computational resources to search our entire training dataset, this could have contributed to our results [29].

It is notable that our best-performing zero-shot Llama2 model was the date-in-context prompt (experiment 4). The task in this study involves temporal relation – identifying the amount of time that has passed from when a seizure occurred to when the note was written – which is a known challenge for existing generative language models[10,13,14,30]. Recent work has shown that GPTs struggle to perform logical reasoning tasks, and rather rely on token bias, or memorized relationships, to correctly answer reasoning problems[31]. While other studies demonstrate that large language models can be taught to learn temporal relation either through prompt engineering, fine-tuning[14], or the introduction of a temporal graph[32], a fine-tuned BERT variant was the highest performing model on temporal relation tasks compared to prompted and fine-tuned GPTs[14]. Deng et al., demonstrated that their C.L.E.A.R. prompt engineering approach can improve parsing through temporal relationships without fine-tuning model parameters [13]. We posit that our “date formatting” instructions, which introduces clear instructions for date extraction and temporal relation procedure like the C.L.E.A.R. method, in conjunction with date-in-context model response fine-tuning can be a mechanism to help GPTs approximate temporal relations without directly addressing token bias.

An important finding of this study was that Llama2 with date-in-context prompts performed worse than fine-tuned Bio+ClinicalBERT on notes written by epileptologists (the training corpus) but *better* than Bio+ClinicalBERT on notes written by neurologists who were not epilepsy specialists. This suggests a tradeoff: fine-tuning on notes written by specialists may improve a model’s performance on that dataset at the expense of generalizability to notes written by non-specialists. Our neurologist notes contained sparser target information (pertaining to seizure freedom) and likely used slightly different and more variable phrasing to describe seizure outcomes. The drop in performance from specialist to non-specialist notes with the fine-tuned Bio+ClinicalBERT model is consistent with previous studies showing that BERT models can fail to generalize to new contexts with different underlying distributions[33], and is well documented in our previous work[9]. Our zero-shot Llama2 model performed better in the sparser context, and similarly between specialists and non-specialists, likely due to the broader domain used during model training[20], and the large number of parameters enabling complex text extraction[34]. However, this generalizability was not unlimited: both models performed poorly on notes written by non-neurologists.

Although our best-performing Llama2 model underperformed the fine-tuned Bio+ClinicalBERT model, we wondered if the difference in performance would lead to meaningful differences in outcomes when deploying the models in real-world clinical research. We replicated a previous analysis of health disparities between demographic groups[3] and found similar conclusions using both models. This indicates that even a less-than-optimal language model can produce meaningful results in real-world applications and may argue for zero-shot approaches that require less initial human effort to implement. Regarding the discrepancy in extracting seizure freedom based on sex: the cause is not immediately clear and will be the subject of future research. However, we previously demonstrated that the Bio+ClinicalBERT model had no differences in extracting seizure freedom between male and female patients. Here we observed that Llama2 systematically under-predicted seizure freedom for female patients (or over-predicted seizure freedom in male patients). Sex-based bias is a known issue for GPT-generated text[35–37]. We found that, in our context, Llama2 interprets clinical notes and generates outcomes differently for men and women – information that is frequently explicitly or implicitly included in medical notes. Future work should continue mitigating GPT bias both during training and in post-hoc analyses of their predictions[38,39], improving both performance and reliability of studies leveraging GPTs. Our findings ultimately emphasize that outcomes derived from Llama2 with date-in-context prompting in a zero-shot setting can be used to perform large scale population studies.

In addition to demonstrating that we can use GPTs to reliably extract seizure freedom in a variety of clinical and research settings, we also provide a framework for future researchers to leverage GPTs for EHR studies. Previous efforts in our group to extract seizure freedom[4], seizure frequency, medications[40], and clinical metadata have relied on manually defined rule-based extraction, or fine-tuning Bio+Clinical_BERT models to accomplish specific extraction tasks – which requires manual annotation of clinical notes. Other recent work has leveraged manual NLP methods to extract stroke severity[41], infectious disease symptoms[42], opioid use[43], among other use-cases, while other efforts demonstrate high performance in information extraction but require annotated clinical notes for fine-tuning. Yan et al. demonstrated the ability of GPT-3.5 and GPT-4 to create SQL queries that parse through the EHR, in part reducing the need for expert input in generating rules-based algorithms[44]. However, this method still requires practitioners to have sufficient background to implement the algorithms. Our GPT pipeline for outcome extraction includes example instruction fine-tuning prompt formats, and scripts that can be easily adopted by new practitioners with limited knowledge of databases or data science, and significantly reduces the clinical burden of manual note annotation and rules-based algorithm generation for future work. In the future, we plan to apply the prompting methods we develop here to extract and study potential risk factors for a variety of episodic diseases and patient outcomes. Rapidly deploying zero-shot GPT pipelines for EHR text extraction can improve early screening and help identify effective treatments.

Our best-performing date-in-context prompt had shorter inference time compared to several other prompt engineering strategies. Recent examinations of instruction fine-tuning have demonstrated that including training examples in prompts teaches the model how to format answers rather than how to answer correctly[28]. We hypothesize the decrease in inference time is explained by our brief, one-sentence synthetic GPT responses in the date-in-context prompt - that inference time was influenced more by the length of the output, rather than the length of the input. The formatting effect of instruction fine-tuning is further demonstrated in our instruction fine-tuning experiments, where we saw no significant decrease in inference time when adding three training examples even though the prompt length substantially increases. Furthermore, the general decrease in performance with the addition of instruction fine-tuning suggests that no additional learning is occurring, which has been demonstrated in other EHR studies[28,45]. Through date-in-context, we demonstrated an effective prompt that attempts to maximize the tradeoff between performance and inference time that can be used to extract complex outcomes in episodic disorders in a completely zero-shot setting.

Our study had several limitations. Generating predictions using Llama2 13B requires significant computational resources that present a computational burden on the user as well as an environmental burden during the training and inference process. Generating predictions locally on 10,000 notes would take approximately 30 hours with our best GPT prompt, but only 13 minutes with our BERT model. While this is currently a limitation, smaller and more powerful models[46–48] reduce the need for computational power and engineering advancements reduce the environmental impact of generative AI[49]. Furthermore, there is an emerging trend of research institutions offering HIPAA compliant access to the largest GPTs[50–52], which could enable researchers to utilize even more capable models and larger context windows to study the EHR at scale. Deep learning, LLMs, and especially GPTs are all relatively black box models[53,54]: very little is understood about the actual computational process that generates each new token. Prompt engineering as a science is even more of a “black box” field as it is both nascent, arriving with the wide-spread access to ChatGPT[55], and sensitive – small prompt variations can cause large changes in model output[56]. Further work is needed to better understand how prompt engineering can reliably increase performance in text extraction and temporal reasoning tasks on the EHR.

In conclusion, the fine-tuned Bio+ClinicalBERT model outperformed the zero-shot Llama2 model in the presence of large amounts of manually annotated notes. Despite this, Llama2 was able to replicate a real-world analysis and generalized better to nonspecialist notes, without the need for manual annotations in a zero-shot setting. Different prompt engineering strategies produced very different outcomes, and the optimal prompt engineering strategy for temporal reasoning tasks is an important area of future research. Continued research into the zero-shot and few-shot application of GPTs to extracting clinical outcomes and risk factors will enhance our ability to leverage the large amount electronic health data available globally and ultimately improve patient outcomes through a revolutionized clinical experience.

## Data Availability

All data produced in the present study are unavailable to protect patient privacy.

## 5. Data and Code availability

We do not make our data available to the public to protect patient privacy. Our fine-tuned NLP models are available on the Hugging Face hub at https://huggingface.co/CNT-UPenn and our code will be made available on GitHub at https://github.com/wojemann/GPT-seizure-freedom.

## 6. Acknowledgements

We would like to thank Alfredo Lucas for valuable discussions during project ideation.

## 7. Funding Declaration

This research was funded by the National Institute of Neurological Disorders and Stroke DP1NS122038; by the National Institutes of Health R01NS125137; the Mirowski Family Foundation; by contributions from Neil and Barbara Smit; and by contributions from Jonathan and Bonnie Rothberg. W.K.S.O. was supported by the National Science Foundation Research Grant Fellowship DGE-1845298. C.A.E. was supported by the National Institute of Neurological Disorders and Stroke of the National Institutes of Health Award Number K23NS121520. D.R.’s work was partially funded by the Office of Naval Research Contract N00014-19-1-2620.

## 8. Author contributions

**WO:** Conceptualization, Methodology, Software, Validation, Formal analysis, Writing – Original Draft, Writing – Review & Editing, Visualization, Supervision, Project administration. **KX:** Conceptualization, Methodology, Validation, Formal analysis, Investigation, Data Curation, Writing – Original Draft, Writing – Review & Editing, Visualization. **KL:** Methodology, Software, Investigation, Data Curation, Writing – Original Draft, Writing – Review & Editing, Visualization. **EC:** Writing – Original Draft, Writing – Review & Editing. **DR:** Writing – Review & Editing, Supervision. **BL:** Resources, Writing – Review & Editing, Funding acquisition. **CE**: Conceptualization, Data Curation, Writing – Original Draft, Writing – Review & Editing, Funding acquisition.

## Notes

### Competing Interest Statement

The authors have declared no competing interest.

### Author Declarations

The IRB of the University of Pennsylvania waived ethical approval for this work.

